# Health care worker’s perspective on the MDR-TB treatment guidelines in South Africa

**DOI:** 10.1101/2024.11.09.24317027

**Authors:** Serisha Ramasir, Frasia Oosthuizen, Varsha Bangalee

**Affiliations:** University of KwaZulu Natal

## Abstract

**Background:** Treatment guidelines are developed and updated according to evidence-based research. Healthcare workers in the public health sector are responsible for the implementation of said guidelines. The multi-drug resistant tuberculosis (MDR-TB) guideline has undergone major changes over the past 5 years. These changes included shorter treatment regimens and the introduction of novel drugs. These changes have consequently impacted the management of MDR-TB patients.

**Objectives:** To establish healthcare workers’ perspectives on the MDR-TB guidelines and managing patients with MDR-TB.

**Methods:** Healthcare workers employed in MDR-TB departments at Murchison District Hospital in KwaZulu-Natal were invited to complete a written questionnaire. These healthcare workers comprised of doctors, nurses, pharmacists, and data capturers. The questionnaire consisted of both open- and closed-ended questions. Descriptive statistics were generated for closed-ended questions using SPSS (version 25). Open-ended questions were transcribed onto Microsoft Excel and analyzed for common and emerging themes.

**Results:** There were 44 staff employed in the inpatient and outpatient MDR-TB unit. Thirty-four of participants agreed to participate in the questionnaire. HIV co-infection was highlighted as a contributor to complicating the management of MDR-TB patients. The most common co-morbidities associated with MDR-TB patients were diabetes, anaemia and hypertension. The socioeconomic influence on outcomes were highlighted throughout patient responses.

**Conclusion:** The MDR-TB program has come been progressive in its approach to gaining control over MDR-TB. The SBR is an effective regimen with a few safety concerns. The nature of ADRs associated with the regimen are serious with myelosuppression highlighted as a common ADR. The impact of socio-economic factors on treatment outcomes is an important factor that should be considered in addition to MDR-TB regimens. The BPaL regimen may offer benefits of a shorter duration and reduced pill burden but there are concerns over the safety of the regimen. The success of the MDR-TB program requires a holistic approach combining patient factors with an effective, safe and convenient regimen.

## Introduction

Multi-drug resistant tuberculosis (MDR-TB) is a global health crisis that is attributed to the evolution of *Mycobacterium tuberculosis* bacteria that has developed resistance to first-line drugs used in the treatment of drug-sensitive TB (DS-TB) i.e., rifampicin and isoniazid. The problem is fueled by the misuse of treatment, mismanagement of MDR-TB drugs through incorrect dosages, drug interactions, and largely through poor patient adherence. This combination of factors as well as the continued transmission of resistant bacteria results in the growing global crisis experienced currently (1,3).

The association between TB and poverty is well documented (21). This is further illustrated by the distribution of TB infections globally. The worst affected countries include middle to low-income countries with poor socio-economic conditions. Access to healthcare, medication, food, and proper housing with sanitation are all additional factors that impact directly on patient adherence and outcomes. Such factors are often overlooked when considering strategies for tackling the TB crisis. South Africa ranks among the top 10 highest TB-burdened countries. According to the 2020 cohort, South Africa had 6784 cases of drug-resistant TB. The WHO monitors and reports on the countries with the highest burden for TB, HIV-associated TB, and MDR/RR-TB. South Africa is no exception to the impacts of TB as it appears in all these reports (5,7).

The treatment of MDR-TB was historically known to be a lengthy regimen associated with poor outcomes and a multitude of adverse drug reactions (ADR) (1,2). The injectable-based regimen that was used prior to 2018, was associated with low treatment success rates of 55%. Since then, the development of two new drugs, bedaquiline, and delaminid, led to a new era in MDR-TB management. The South African National TB program (NTP) used data from the Standard Treatment Regimen of Anti-tuberculosis Drugs for Patients With MDR-TB (STREAM) Trials and the well-documented “Bangladesh regimen” to implement a standardized approach to MDR-TB treatment. The data from these pilot studies demonstrated improved outcomes with a better safety profile compared to the 18-24 month injectable-based regimen used previously in SA. The new regimen termed the shot bedaquiline regimen (SBR), incorporated bedaquiline and two new re-purposed drugs, clofazimine and linezolid, and boasted a shorter treatment duration of 9-11 months (4, 6).

In South Africa, bedaquiline was introduced via a phased programmatic approach i.e. The Bedaquiline Compassionate Access Program (BCAP). This program was introduced in 2013 and limited the use of bedaquiline to specific hospitals that screened patients and initiated them on bedaquiline-containing MDR-TB regimens per strict inclusion criteria. This controlled use of bedaquiline yielded high rates of treatment success. This was followed by the recommendation that bedaquiline was to be used for patients experiencing toxicity to kanamycin in an 18–24-month regimen. In 2019, the interim clinical guideline was released detailing the use of bedaquiline as part of a standardized regimen for the treatment of uncomplicated MDR-TB. The standardized short bedaquiline regimen (SBR) was 9-11 months long and comprised of a 4–6-month intensive phase and a 5-month continuation phase (23,24).

A successful TB program’s core component would be to provide a safe and efficacious regimen. The World Health Organization (WHO) reported that the treatment success rate from the 2018 cohort was 59% as documented in the global TB report. This was showing steady progress from previous years (25). This is still far from the goals highlighted by the WHO which strives for a treatment success rate of 90%. A new regimen is proposed for use. This new regimen, is six months in duration and consists of four drugs. The regimen comprises of bedaquiline, linezolid, new drug, pretomanid and levofloxacin (BPaLL). This shorter, all-oral regimen will also provide a reduced pill burden compared to current regimens.

The Ugu district is located on the South Coast of KwaZulu Natal. In the district, HIV and TB contribute to 35% of mortality, more than any other single disease or infection). Before the decentralization of MDR-TB services, Murchison hospital was the only hospital in the district providing care to patients diagnosed with MDR-TB. The decentralization of MDR-TB services has since resulted in either patients being initiated at Murchison Hospital and down-referred for the continuation of care, or a small cohort being initiated at approved sites. Murchison hospital remains the primary site for treatment, having two inpatient wards for admission of MDR-TB patients and an MDR-out patient department. The unit has a multidisciplinary team comprising of nurses, doctors, social workers, pharmacists, TB tracers, and administration staff. The MDR-TB guidelines in South Africa are in constant revision in line with updated information from the WHO. The implementation of the guideline requires the combined efforts of medical, nursing, and social services. These frontline participants are key to providing insight into the social and economic factors that directly impact patient outcomes. They may highlight important information on the advantages and disadvantages of regimens used and provide views on possible new regimens (7,26). These views had not been explored previously but may provide holistic insight into the MDR-TB program.

The study aimed to look at the functionality of the MDR-TB program from the viewpoint of those who are implementing the guideline. This would include assessing the roles and responsibilities of participants, establishing their knowledge of the guidelines, assessing the impact of socio-economic factors and HIV as a primary co-morbidity, and holistically establishing a patient profile for a typical patient with MDR-TB in the Ugu district. The treatment regimens used historically and currently will be analyzed in terms of their success as perceived by these participants. The aspects on safety and efficacy will be sought. These findings would provide useful insight into the factors not considered or often overlooked when guidelines are revised and may also be useful in creating a more integrated MDR-TB guideline.

## Methods

### Study design and population

The study was conducted between the period 28 June till 2 September 2021. Printed questionnaires were handed out to all members of the study population. The study population consisted of healthcare workers such as doctors, nurses, pharmacists, pharmacist assistants, data capturers and TB tracing teams.

Purposive sampling was used in the recruitment of participants. This was the most appropriate sampling strategy considering the small sample size and nature of the questionnaire (9). Participants were invited to a meeting that provided information about the study and for the recruitment of interested participants. They were allowed to seek clarity on any areas of uncertainty. The timeline for completion was also communicated at this meeting.

The sample size was calculated using an online sample size calculator, Surveymonkey®. The eligible population at Murchison Hospital was found to be 40 participants at the point of data collection, therefore a minimum number of 37 participants were required to achieve a 95% confidence interval.

### Ethical clearance

Ethical clearance to conduct the study was obtained from the Biomedical Research Ethics Committee at the University of KwaZulu-Natal, Durban (BREC/00002065/2020). Gatekeeper approval was obtained from the KwaZulu Natal Department of Health as well as the hospital where the study was undertaken.

### Study Questionnaire

The study utilized a manually distributed written questionnaire after considering the challenges of connectivity and computer literacy of participants.

The questionnaire was designed to consider the objectives of the study and the information required to ascertain this knowledge. There was consideration given to the fact that the questionnaire would be completed by various categories of healthcare workers therefore it was designed to be inclusive of all these categories with respect to clinical literacy.

The questionnaire was divided into three sections comprising of 29 open and closed-ended questions. Section A comprised of six questions and aimed at assessing the role of the participant in the healthcare chain and their knowledge and training of MDR-TB guidelines and treatment. Section B included questions that were based on the socioeconomic factors influencing patient outcomes, the co-morbidities commonly encountered in patients in the district and their impact on treatment, and an overview of the TB program. The questions in this section were aimed at ascertaining information on practical experiences regarding the management of patients with MDR-TB as well as establishing their views on the safety and efficacy of regimens. Section C comprised of questions on the MDR-TB regimens and their safety and efficacy.

The questionnaire was piloted amongst seven individuals at a different hospital to where the study was conducted. This was done to ensure the questions were understandable, clear, and logical. Minor amendments were made as a result of this exercise. The questionnaire was given final approval by two senior academic pharmacists at the University of KwaZulu-Natal.

### Data analysis

All responses were captured on an Microsoft Excel® spreadsheet that differentiated open and closed ended questions. The responses were analyzed by thematic analysis for open-ended, qualitative responses. Close-ended questions were analyzed descriptively using SPSS (version 27).

## Results

There was a response rate of 85% (n=34). All questionnaires were assessed for completeness on collection. There was a minimal number of missing responses (n=32) and these pertained mainly to open-ended questions where justification of responses was required.

The total number of participants consisted of 31 clinical and 3 non-clinical participants. There were22 nurses, 5 medical doctors, 3 pharmacists and 1 pharmacist assistant. There was 1 administration clerk, 1 data capturer, 1 outreach service staff in the non-clinical component

### Section A: Knowledge and training on MDR-TB guidelines

In terms of knowledge of the MDR-TB guideline and updates to the guideline, 28 participants expressed confidence in their skills. The remaining six felt unsure of their skills and were found to be lacking in their knowledge of MDR-TB guidelines. These consisted of both clinical and non-clinical participants.

Eight participants further expressed that they had not received adequate training on guidelines and changes to the guidelines while the remaining twenty-six felt the training provided was adequate and felt confident in their knowledge of medication, interactions, and side effects. This group was provided this knowledge and skills in the form of practical training in the workplace from colleagues. Formal structured training was provided to two participants. Those participants who had identified a training need requested that this be done in a formal and structured manner

### Section B: TB program/Co-morbidities/Socioeconomic factors

The questions in section B were posed to determine the perceptions on the relationship between MDR-TB and factors such as co-morbidities, socioeconomic factors, adherence, monitoring, regimen design, and resistance development based on participants’ practical experiences. Responses to the closed-ended questions are presented in table 1 below.

**Table 1:**
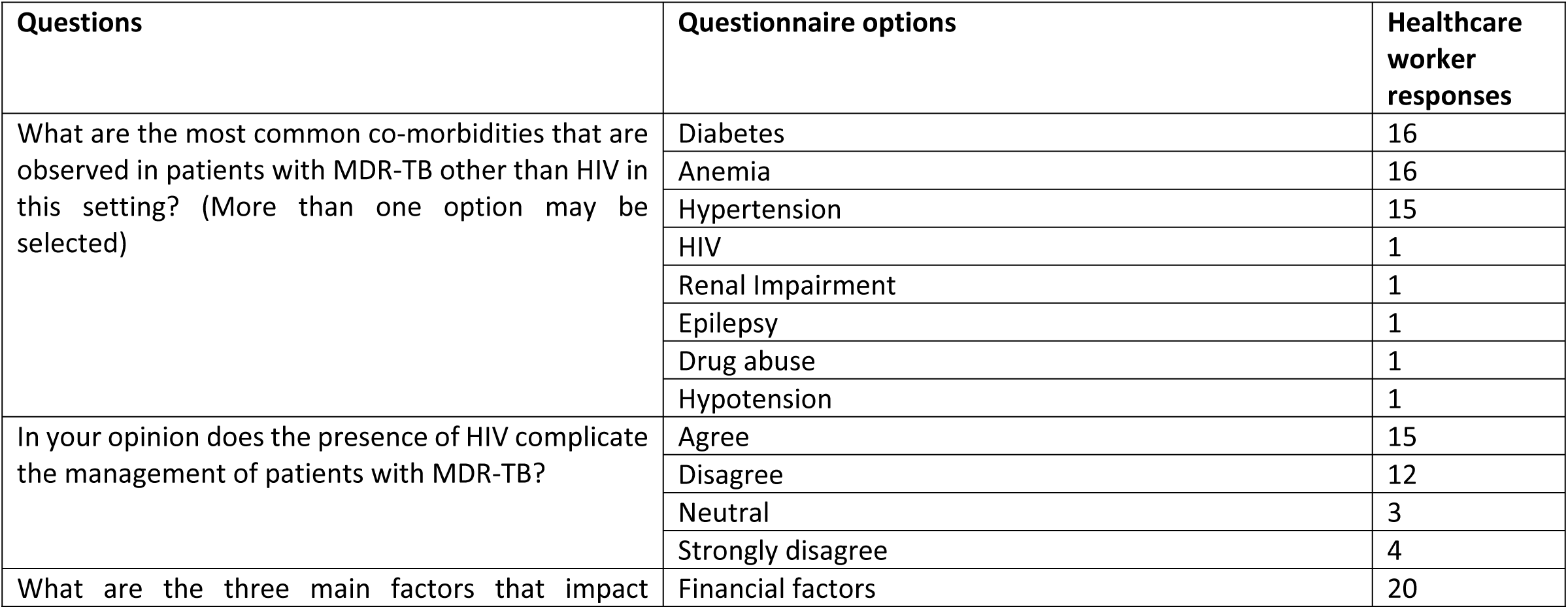
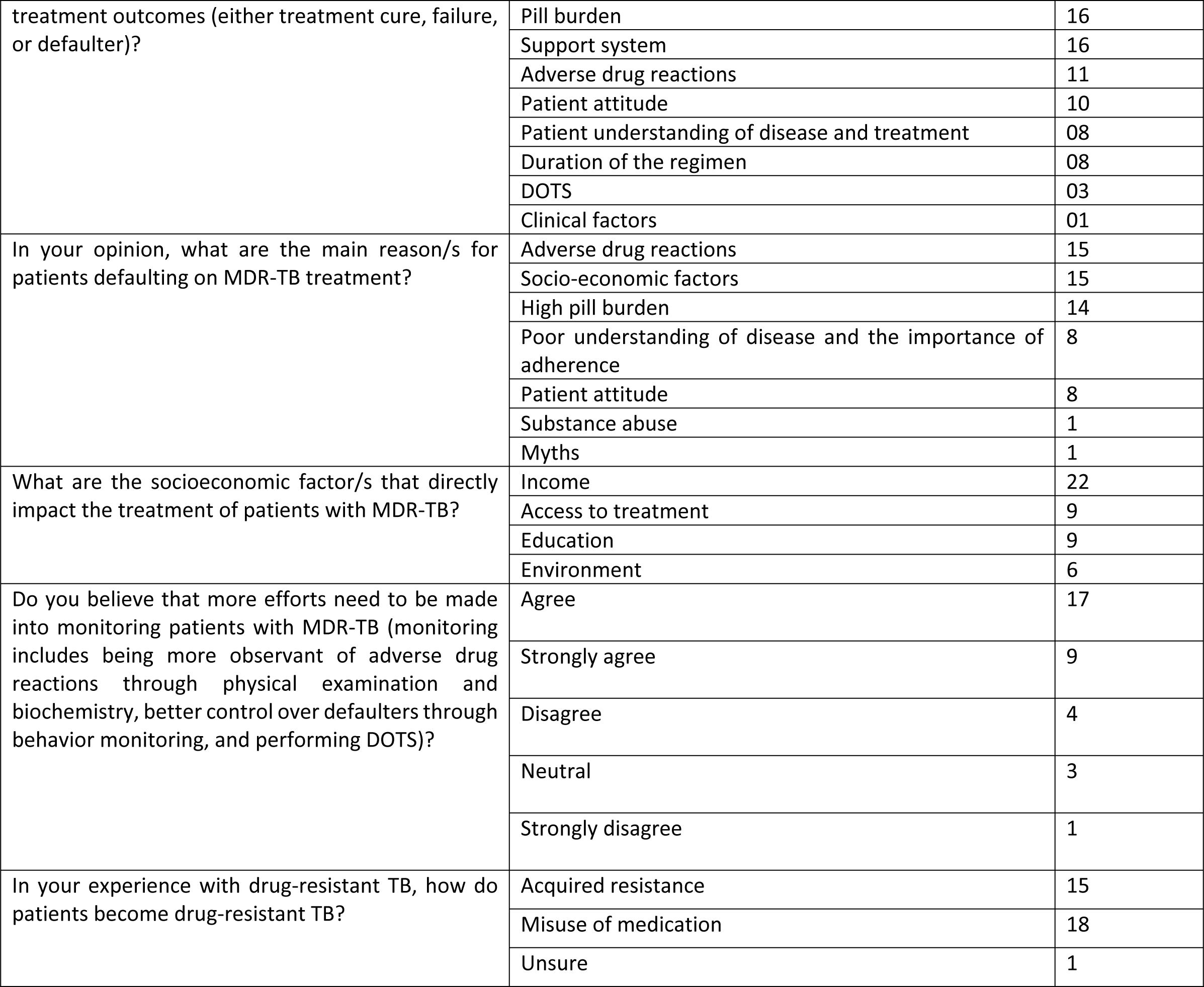
representation of results of closed-ended questions for section B.

Section B consisted of 4 open-ended questions. The emerging themes are presented in table two.

**Table two.**
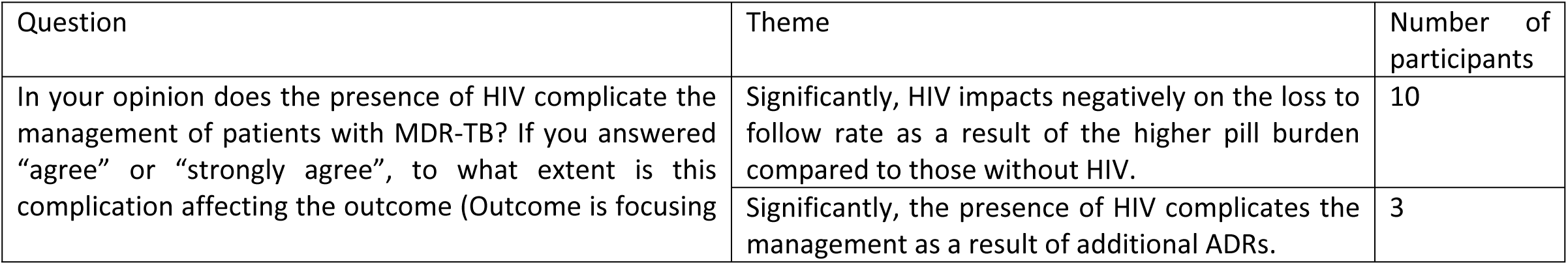
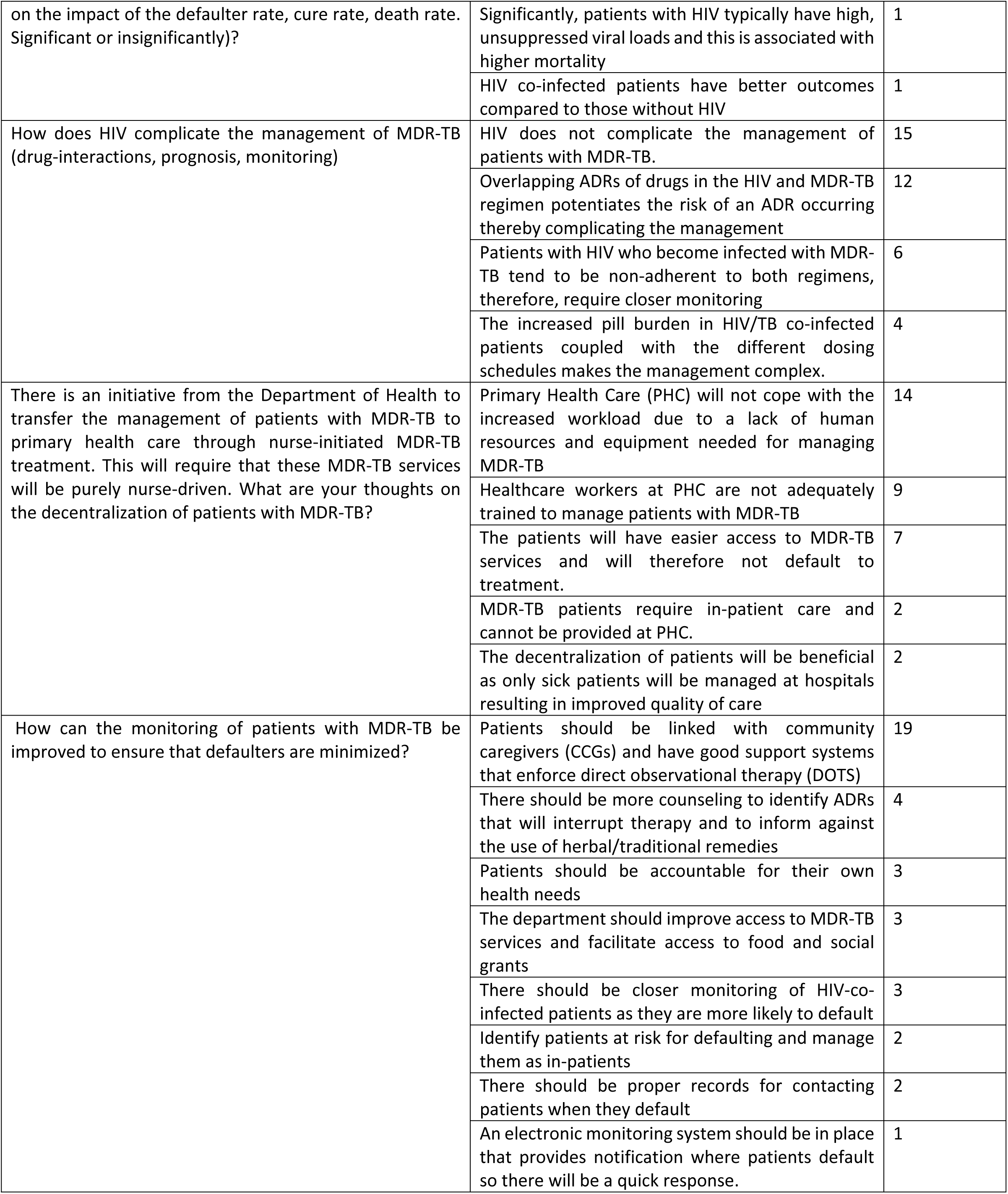
represents the thematic responses to open ended questions in Section B.

### Section C: MDR TB regimens

This section comprises of 22 questions (6 open-ended questions and 16 closed-ended questions). This section focused on the different regimens used to treat MDR-TB. It aimed to compare the different regimens with respect to their safety and efficacy. The section also explored future treatments and healthcare worker perceptions regarding this. The closed-ended questions are represented in table three and the themes from open-ended questions are reflected in table four

**Table three.**
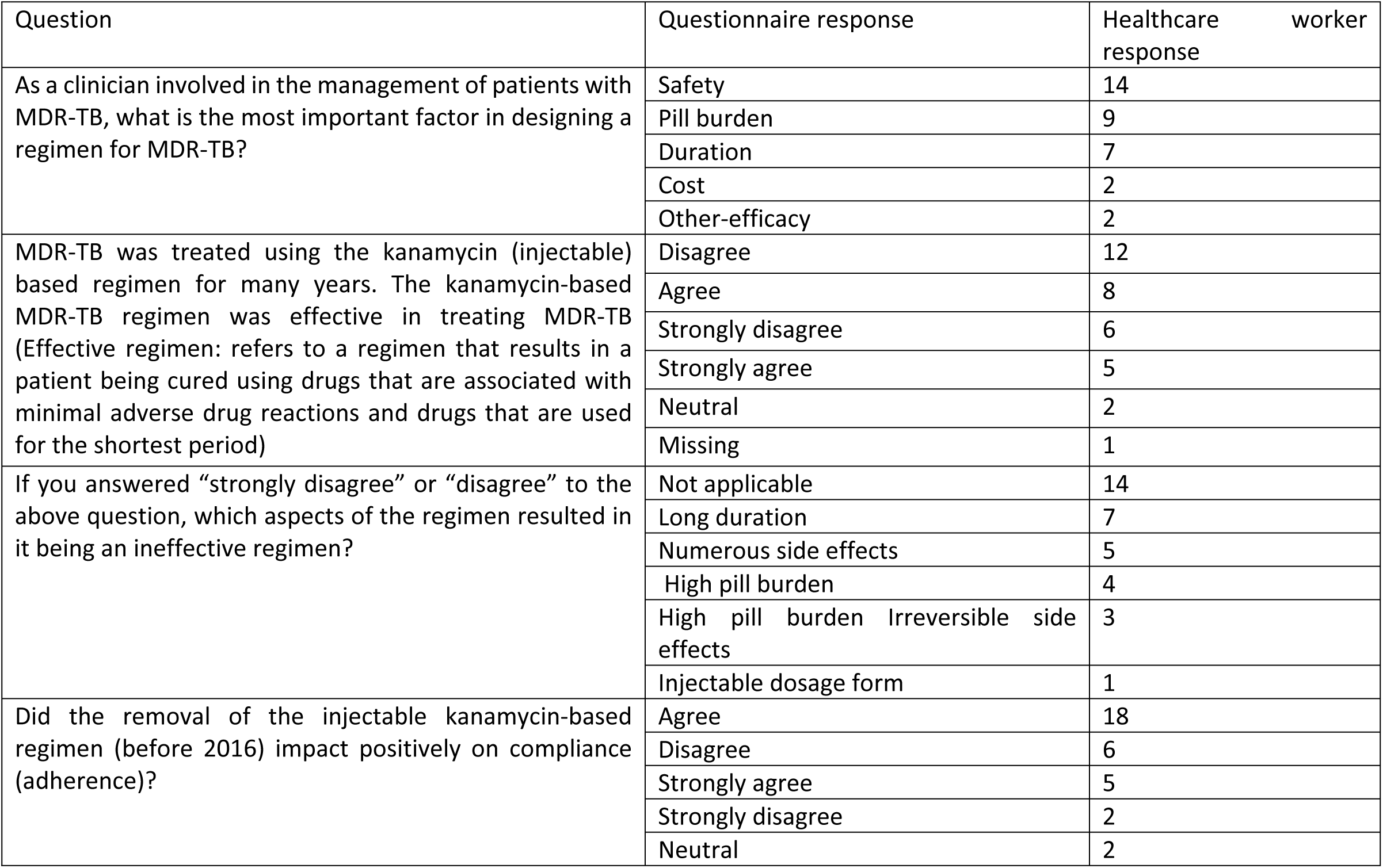
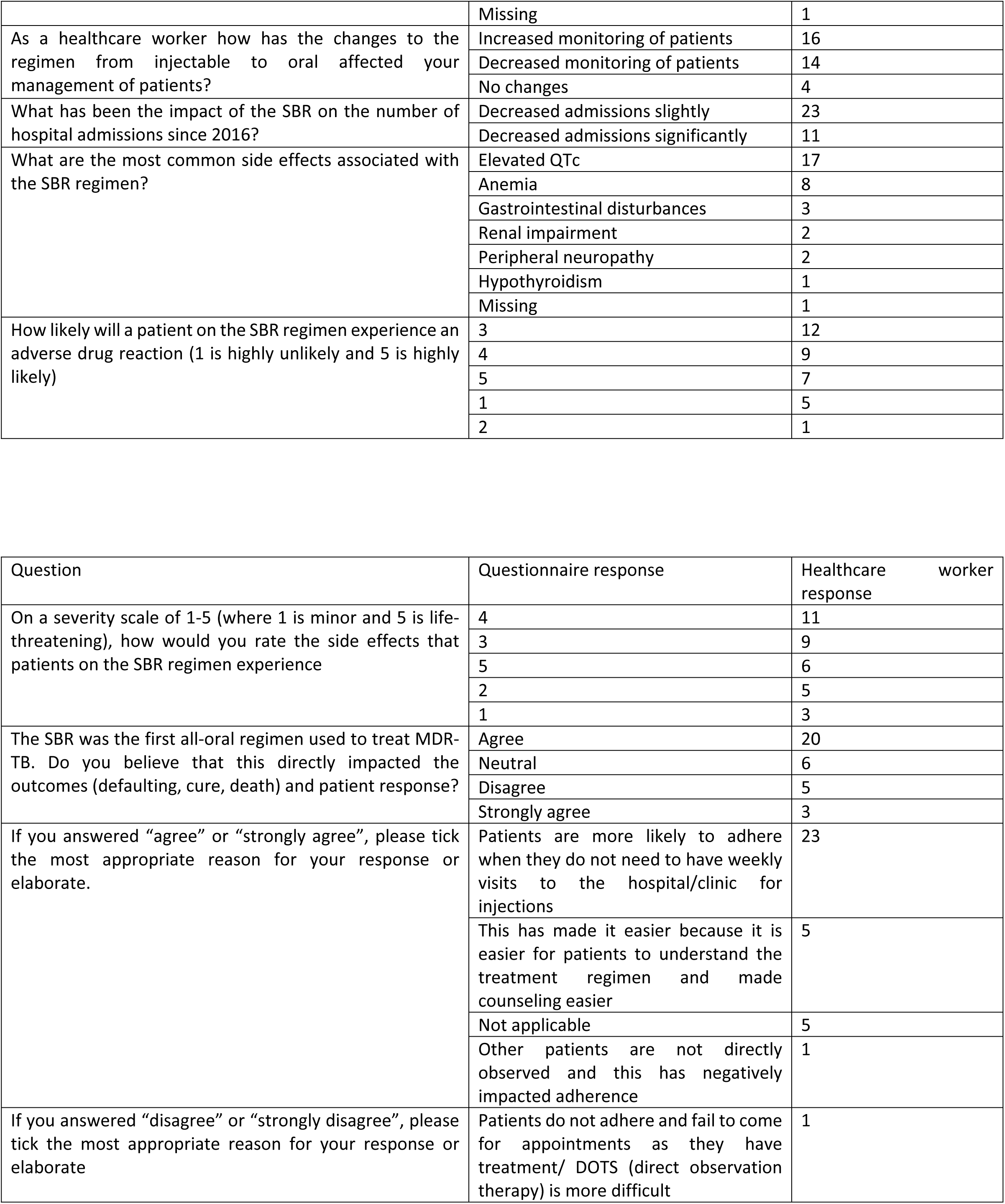
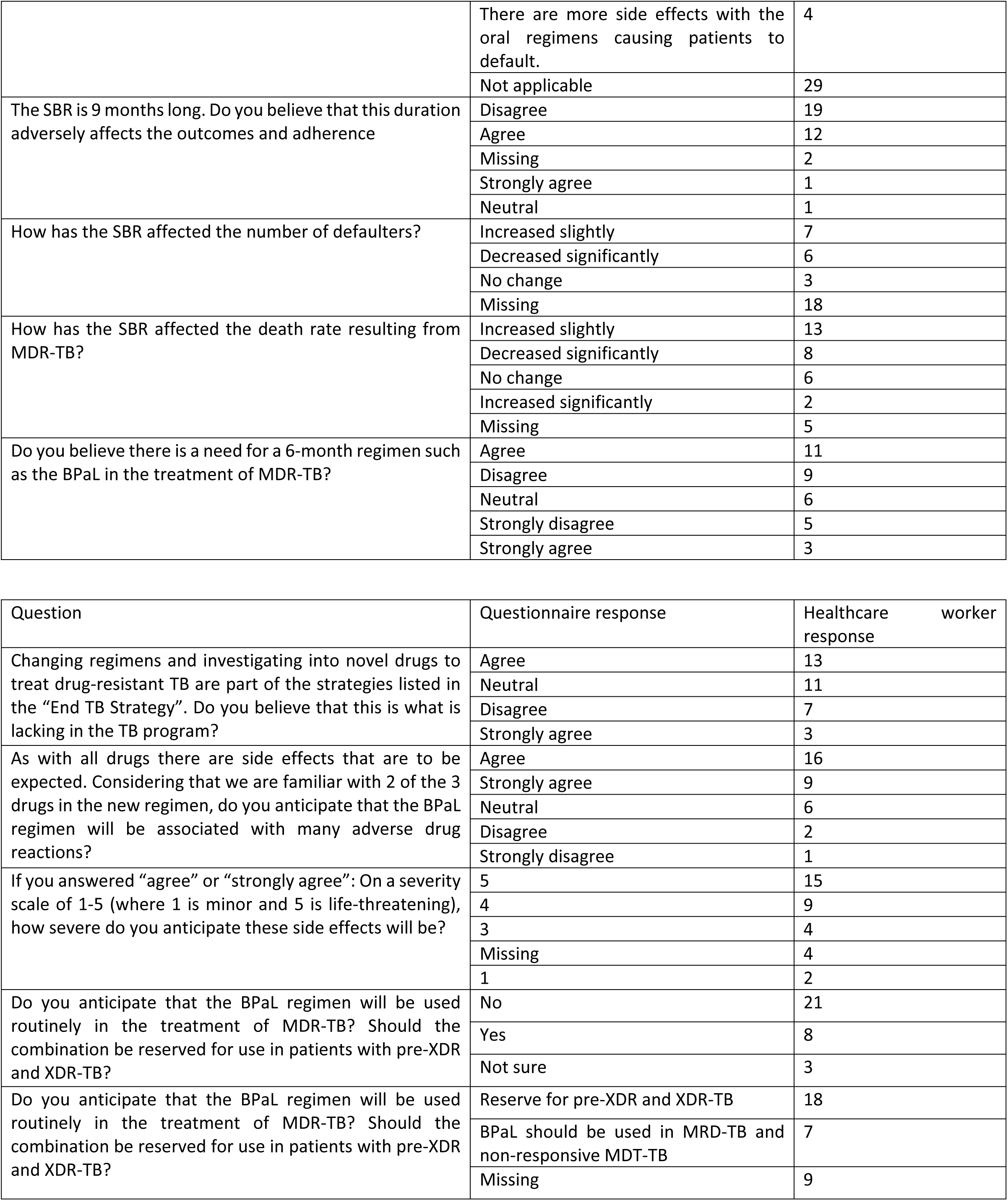
represents the results of enclosed-ended questions from Section C.

**Table four.** shows the thematic responses to open-ended ended questions in Section C.

| Question | Theme | Number of participants |
| --- | --- | --- |
| Did the removal of the injectable kanamycin-based regimen (before 2016) impact positively on compliance (adherence)? Please elaborate | Missing | 23 |
|  | The kanamycin-based regimen is associated with ADRs | 4 |
|  | The kanamycin-based regimen is lengthy and this causes a high loss to follow rate | 3 |
|  | The SBR is associated with higher mortality compared to the kanamycin-based regimen | 3 |
|  | Socio-economic factors impact compliance to a greater extent than the regimen used | 1 |
| What aspects of the SBR do you find more useful? | The SBR is a shorter regimen | 11 |
|  | The SBR is associated with a better side effect profile | 6 |
|  | The all-oral regimen is more convenient for patients | 5 |
|  | None | 4 |
|  | The SBR has a reduced pill burden compared to the kanamycin-based regimen | 3 |
|  | There is fewer loss to follow patients compared to the kanamycin-based regimen | 2 |
|  | There is a faster clinical response in patients treated with the SBR ie. Faster sputum conversion rate | 1 |
|  | The SBR is not more useful compared to the kanamycin-based regimen as it has a worse side effect profile | 1 |
| What aspects of the SBR do you find less useful? | There are a greater number of occurrences of ADRs associated with the SBR | 15 |
|  | The ADRs associated with the SBR is more severe compared to the ADRs associated with the kanamycin-based regimen | 5 |
|  | There are no disadvantages associated with the SBR | 5 |
|  | There is an increased number of loss to follow patients associated with the SBR | 4 |
|  | There is more monitoring required in patients using the SBR | 3 |
|  | There is an increased pill burden compared to the kanamycin-based regimen | 2 |
| Changing regimens and investigation into novel drugs to treat drug-resistant TB are part of the strategies listed in the “End TB Strategy”. Do you believe that this is what is lacking in the TB program? | Missing | 27 |
|  | No, there should be more focus should be on social determinants of health | 4 |
|  | No, there is no problem with access to efficacious medication as the treatment that is available is sufficient | 2 |
|  | No, there should be greater emphasis on TB preventative strategies | 1 |
| Do you anticipate that the BPaL regimen will display a greater number of positive outcomes compared to the SBR if it is routinely used in the treatment of MDR-TB? Please elaborate | Missing | 13 |
|  | The shorter duration of the BPaL will lead to fewer losses to follow patients | 5 |
|  | The BPaL regimen is anticipated to have more severe ADRs | 4 |
|  | The current regimen design is sufficient for achieving positive outcomes and there is no need for additional regimens | 3 |
|  | The shorter duration of the BPaL regimen will not significantly reduce the loss to follow | 3 |
|  | There is a need for more information on the safety and efficacy of the BPaL regimen | 2 |
|  | The use of novel drugs should be limited to prevent resistance | 2 |
|  | The number of ADRs will be reduced if the BPaL regimen is used | 1 |
|  | The SBR is associated with poor outcomes | 1 |

## Discussion

The purpose of the study was to explore the perceptions of healthcare workers on the MDR-TB program and factors impacting the outcomes and the safety and efficacy of MDR-TB regimens

The majority of participants were nurses. This is reflective of the majority of public sector programs where nurses are the center of patient care, management, and follow-up.

*Singh et al.* aimed to establish the knowledge of nurses on MDR-TB. This was done in anticipation of the decentralization of services to primary health care (PHC) in the Eastern Cape province. This study found that there was a gap in the knowledge on how to manage MDR-TB associated ADRs. This could have implications for patient adherence and subsequent outcomes (31). The questionnaire conducted currently did not seek specific areas of uncertainty however twenty-six participants in the MDR-TB department felt confident in their skills and knowledge of the MDR-TB treatment and guidelines. *Kansal et al*. and *Minnery et al.* conducted studies in India and Peru respectively and these showed that knowledge gaps existed among participants. The consequences of this lack of knowledge results in inadequate treatment (28,30). Both of the studies highlighted the need for adequate training of staff. The decentralization of MDR-TB patients reflected that this initiative requires adequately trained staff and the program would suffer due to lack of resources in resource-limited settings such as PHC. The lack of formal training was highlighted in this study. A formal structured, continuous training program is advisable to ensure that all staff is trained with up-to-date information. This would also provide a solution to high staff turnover and rotations as highlighted by *Loveday et al.* (29). It is apparent that no formal training is provided on the management of patients with MDR-TB and a more practical approach is adopted in this setting.

Aspects that related to MDR-TB disease revealed that the most commonly observed co-morbidities were diabetes, hypertension, and anemia. These were in addition to HIV. These results are consistent with results from *Tao et al*. that found diabetes, hypertension, and chronic obstructive pulmonary disease (COPD) to be common comorbidities that occur with tuberculosis (10). This is important as it highlights a need for closer monitoring of these patient groups for early detection and treatment. The inclusion of anemia on this list is an atypical comorbidity not commonly observed. This may be a complication of HIV treatment or malnutrition due to poor social situations which are both scenarios that are common to the rural South African setting.

Participants expressed that HIV does not complicate the management of patients. Sixteen participants expressed that the presence of HIV is not an additional complication to the management of patients whilst 15 responded that HIV creates an additional barrier to the management of MDR-TB patients. A study conducted by *Elliot et al.* found that the presence of HIV was not associated with poor outcomes (32). The minority who mentioned that HIV complicated treatment management attributed the complication to the increased pill burden in patients with HIV co-infection.

The increased pill burden impacts on compliance resulting in a high number of loss to follows. *Deshmukh et al*. conducted a qualitative study in India to establish reasons for loss to follow. According to the study the pill burden, treatment duration, ADRs, and lack of social support were noted to contribute to loss to follow. This enforces that a higher pill burden caused by taking both antiretrovirals (ART) and MDR-TB treatment contributes to high loss to follow (33). *Amuha et al.* also re-iterated that treatment for HIV and MDR-TB results in high pill burdens which are directly linked to non-adherence (11).

Participants noted that there are more ADRs in patients with HIV co-infection and the management of these ADRs holistically complicates the treatment. HIV-positive patients with CD4 counts <100 cells/mm^3^ and those initiating newly initiating ART, are at higher risk for experiencing severe ADRs which in turn contributes to a higher risk of death or loss to follow (34). One healthcare worker felt that those patients who have been on ART and still have high viral loads are often non-adherent and this is an indicator of non-adherence to MDR-TB treatment. Such behavior is a predictor of poor outcomes. This can be useful in identifying those patients at high risk for defaulting and therefore more efforts can be put in place to identify the reasons for non-adherence and provide more social support at the start of the treatment program. This also highlights the important role played by both HIV and TB in influencing the outcomes of each respective infection (35). A vast amount of literature supports that the presence of HIV does influence MDR-TB outcomes negatively albeit not directly and therefore more attention should be given to patients who are co-infected to ensure that they are provided with additional tools and support for successful outcomes of both infections. Integrated care is also vital and it is recommended that patients should be treated using the “one-stop” approach where all health needs are treated by the same doctor and facility on one visit. This would not only provide a better view of the overall health status of the patient but would encourage compliance with the treatment regimen (49). Participants may be of the latter conclusion due to patients treated using a compartmentalized approach, especially with MDR-TB. The treatment of MDR-TB is often viewed as a specialty with the management of the condition treated a separate department. This may provide the impression that HIV does not create a barrier to management and treatment as HIV is managed as a separate infection in another department.

The three main factors that impact treatment outcomes were identified as financial factors, pill burden, and the presence of a support system. Financial factors were ranked as the most common response reflecting the sentiment that MDR-TB is a disease afflicting the impoverished community (21). These factors are well highlighted in the literature as contributors to loss to follow rates and subsequently poor outcomes. A study conducted in Indonesia by *Soeroto et al.* found that a history of previous TB treatment, time of culture conversion >2 months, and malnutrition were the factors determining negative outcomes (12). Clinically these factors may be significant predictors of poor outcomes however in the South African context with the developing health care system, these participants found clinical factors and DOTs to be the least important determinants of outcomes. A lack of financial resources is a norm in South Africa where 45.5% of households rely on social grants as a source of income (37). This impacts access to treatment and the ability to take treatment effectively. There are social grants and sponsorships through non-government organizations which are available and it would be useful if healthcare facilities could facilitate access to these resources. A range of rehabilitation services is available at hospitals to assist in capacitating patients and encouraging the building of skills that may be income-generating. The second factor of pill burden is a well-known contributor to poor treatment outcomes and it is the aim of newer regimens such as the BPaL regimen to combat this issue by providing fewer medications in a shorter regimen. The third factor of support systems is an integral part of care and this can be facilitated by incorporating family or friends in the treatment plan to aid and support patients in complying with regimens. This also includes patients accessing social services which are available at hospitals for discussion on any barriers to adherence. This would help to keep patients engaged throughout the treatment regimen and facilitate a positive attitude to treatment.

Loss to follow is a major contributor to the development of acquired MDR-TB (38). Participants found ADRs and socio-economic factors to be the most common reasons for patients failing to comply with treatment MDR-TB regimens. High pill burdens were again highlighted as the third most common factor. The study by *Lin et al.* highlighted the following factors as contributors to loss to follow ie, individual factors (age, place of residence, lack of finance, level of education, alcohol use, history of non-adherent behavior), treatment support services, extended time between diagnosis and treatment and ADRs. These factors are all-inclusive and were highlighted in responses from participants. The socio-economic factor that was highlighted to be the most influential on outcomes was the lack of financial income. Eight participants responded that access to treatment and eight also responded that education also impacted this. Education refers to the level of education of patients. Socioeconomic factors are often mentioned as contributors to negative treatment outcomes. TB disease is noted to be afflicting poor and underprivileged populations. In South Africa, there are inequalities in socio-economic status. This imbalance translates to inequalities in health, access to healthcare, and quality of care received (14). Lack of finance was highlighted as the most influential socio-economic factor that directly impacts the treatment of patients. The lack therefore will result in high loss to follow rates or unnecessary treatment interruption. In addition, patients may experience adverse drug reactions as a result of taking treatment on an empty stomach which further propagates loss to follow. The misuse of treatment was found to be the most common manner in which TB is spread. This then creates a cyclical pattern of infection, treatment, loss to follow, and re-infection. The South African Social Security Agency (SASSA) provides grants for all patients diagnosed with MDR-TB to alleviate these issues. This should be made accessible for these patients as a means to overcome this bottleneck. Support structures were also identified as a socio-economic factor impacting outcomes. The presence of family or friends aids in the acceptance of the diagnosis and treatment. This is imperative in patients’ understanding of the diagnosis and for adherence support.

MDR-TB treatment is known to cause a multitude of ADRs such as prolonged Qtc, hypokalaemia, hypomagnesemia, acute kidney injury, hepatotoxicity, hypothyroidism, gastrointestinal (GI) disturbances, peripheral neuropathy, psychiatric disorders, dermatologic disorders, ototoxicity, electrolyte imbalance, hepatic dysfunction, seizures, arthralgia and headaches (40, 41). The overlapping of ADRs by drugs used in the regimens further potentiates the likelihood of an ADR occurring (41). Elevated Qtc interval and anemia were established as the most common side effects associated with the SBR. The elevated Qtc is well documented as a side effect of bedaquiline contained in the SBR. This side effect may also be caused by clofazimine or fluoroquinolones also contained in the regimen. The inclusion of all three drugs increases the potential for the side effect to occur. The documentation of anemia has not been a focus of many studies. The implicated drug would be linezolid. The occurrence of such a side effect would be exacerbated by the presence of HIV when patients take zidovudine-based regimens. In practice, this may be a commonly occurring phenomenon prompting the response from participants. Socio-economic factors of malnutrition may also have an impact as pre-existing anemia may be worsened through the use of linezolid. The safety of the regimen was also highlighted as the most important factor in designing a regimen. The safety of the SBR was highlighted as a concern following the initial rollout of the regimen. The particular concerns were the ADRs caused by bedaquiline such as prolonged QTc. The previously used kanamycin-based regimen was associated with severe irreversible ADRs which prompted the changing of the injectable-based regimen to SBR (22). The link between ADRs and compliance to the treatment regimen is well documented and its influence on establishing a suitable standardized regimen. This highlights the need for pharmacovigilance. The monitoring of drug safety in the TB program is known to be sub-optimal. The implementation of active TB drug safety monitoring and management (aDSM) was seen as a necessity. aDSM comprises of an active and systematic assessment of patients on treatment with new TB medicines, or novel MDR-TB or XDR-TB regimens, to detect, manage and report suspected or confirmed drug toxicities. It requires that national programs document and report all serious ADRs (42). This is an important tool for achieving control over ADRs and achieving fewer loss to follow but its implementation in the South African setting is found to be lacking. High pill burden and lengthy duration were also highlighted as factors affecting compliance. Participants also identified the importance of considering the pill burden (n=9) and duration of the regimen (n=7) when designing an MDR-TB regimen and its impact on loss to follow. The duration of the regimen was identified as a contributor to high loss to follow rates. This-was not a typical response considering the evolution of the TB program from an 18–24-month regimen to a 9–11-month regimen. This is well documented in various studies and highlights the importance of introducing a regimen such as the SBR that provides the benefits of a shorter regimen with a lower pill burden (14).

The monitoring aspect of the management of patients with MDR-TB is an integral component in the success of patients with MDR-TB. This follow-up care is inclusive of monitoring treatment adherence, monitoring of laboratory markers, counseling on adherence, and identifying and managing adverse drug reactions. 26 participants agreed that more efforts need to be made in this regard. The most commonly suggested way in which this can be improved was the linking of patients with community caregivers (CCGs). Direct observational therapy (DOTS) is a highly effective activity to ensure adherence and compliance to the treatment regimen (15). Participants expressed that increased monitoring of patients is necessary and this would be best achieved through the use of CCGs. These human resources are available and should be utilized in the TB program. They can provide services of conducting home visits as well as assist in monitoring adherence through DOTS. Traditionally patients with adherence issues would have to be admitted to the hospital for DOTS. The reinforcement of CCGs in the TB program can assist in providing these services while patients are managed as outpatients. This can also provide the support system that many patients lack and this may have a positive impact on patient outcomes. The importance of support structures was highlighted by 8 participants. These participants felt that this support should come from family and friends. The involvement of family members or friends as support structures is important to minimize loss to follow and promote patients completing the course of therapy. This would include contact persons if patients fail to adhere to appointments or any family member to accompany patients and provide support in treatment adherence. Proper note-taking of patient details and contacts was highlighted by 2 participants. Counseling on ADRs was identified as a tool to improve the monitoring of patients. This includes counseling on the use of herbal remedies and their potential interactions with treatment. This information can better assist patients in coping with ADRs. It was highlighted that high-risk patients for poor adherence must be inpatients. This is not ideal as part of decongesting healthcare facilities and the extended hospital stay for stable, clinically well patients may be perceived as financially wasteful. Three participants mentioned that no additional initiatives are required as patients should bear more responsibility for achieving positive health outcomes. The initiatives such as the provision of treatment and access to social services were perceived to be sufficient. Conversely, 3 participants felt that more efforts should be made to improve access to MDR-TB services and there should be more social support in proving access to food parcels through non-governmental organizations. The HIV co-infected patients were identified by three participants as a high-risk sub-group who are most likely to be a loss to follow. *Schnippel et al*. found that HIV-positive patients who initiate treatment are at a greater risk for loss to follow. They also identified that the first 6 months of MDR-TB treatment is the period when a severe ADR is most likely to occur. Considering the link between ADRs and outcomes like a loss to follow it would be useful to increase monitoring and counseling of patients during this period (34).

Drug-resistant TB was reported to be the result of misuse of medication. Participants have reported that this is the main way in which MDR-TB infections occur (n=18). Drug-resistant TB occurs in a succession following treatment interruptions or inadequate treatment. This is well documented and supported by the fact that many cases of MDR-TB include re-treatment cases of previous loss to follow cases (43). Increasing new evidence postulates that the continual spread of drug-resistant TB is driven by resistant bacteria exacerbated by poor infection prevention control practices (16,17). The participants’ response may likely be a reflection of the infection profile of the population being represented in the study or more likely be a notion stemming from the fact that the majority of cases of MDR-TB are patients who are re-infected. The re-enforcing of infection prevention control practices is imperative in the South African setting where overcrowding of health care facilities, informal dwellings, overcrowded public transport, and inadequate access to basic sanitation are factors that prompt the growing rates of MDR-TB. The decentralization of MDR-TB services was highlighted as a response to mitigating the spread of MDR-TB at drug facilities but the participants of this questionnaire shared the sentiment that primary health care (PHC) facilities are overburdened currently and the introduction of an additional program is not ideal. The additional training required to manage patients with MDR-TB was highlighted and emphasized that for such a program to be managed as a nurse-driven initiative, adequate training is required. The lack of resources both human and physical was consistently highlighted. The advantage of improved access to healthcare was recognized by a small percentage of participants (n=7).

Thematic responses were extracted from responses to open-ended questions. It was noted that the response to open-ended questions was not consistent compared to closed-ended questions. Participants were reluctant to write responses. These may be due to the inability to express opinions in the preferred language as the majority of the participants did not speak English as a first language. This may also be attributed to the fact that the completion of the questionnaires was done amidst work responsibilities.

The kanamycin-based regimen was well documented to be insufficient in achieving acceptable levels of treatment success rates. The high rates of unsuccessful treatment were attributed to the length of treatment, toxic drug effects, and the high number and low efficacy of standard MDR-TB medications (44). 18 participants agreed with this characterization of the kanamycin-based regimen. The reason provided for these thoughts were also listed. The long treatment duration was the most common response. 14 participants favored the use of the kanamycin-based regimen. The reasons for this were not sought through the questionnaire however participants elaborated on the fact that the SBR is associated with higher mortality. It was reiterated that socioeconomic factors influence outcomes and regimens play a minor role in treatment outcomes. This reflects an uncommon opinion contrary to the vast literature that supports the use of SBR as a more efficacious and safer regimen over the kanamycin-based regimen (16). Based on the majority of responses that favored the SBR, many (n=23) expressed that the kanamycin-based regimen negatively impacted compliance and therefore the removal of that regimen resulted in an improved loss to follow rate. The high number of adverse drug reactions, pill burden, and socioeconomic factors were mentioned to impact compliance. Two of these factors are positively associated with the kanamycin-based regimen.

In the monitoring of patients for adherence, efficacy, and safety laboratory markers, the kanamycin-based regimen focused on monthly audiology examinations, renal function tests, and sputum and culture tests amongst others. The monitoring of patients using the SBR includes tests done with the kanamycin-based regimen but tests such as ECG monitoring, hepatic function, electrolytes, and lipase is essential (45). This implies increased monitoring. 16 participants felt that there is increased monitoring required with the SBR regimen while 14 felt there is decreased monitoring in light of the oral nature of the regimen. Additionally, there should be increased monitoring required for patients using the SBR as it is a relatively new regimen.

The main advantage of the SBR was identified to be the short duration. The main disadvantage was mentioned to be the increased adverse drug reactions. The all-oral regimen was thought to decrease hospitalization as a result of MDR-TB. The absence of injectable treatment meant that patients could be managed as outpatients. Only patients who were acutely ill or experienced a severe ADR may be admitted (17). This decrease is favorable as fewer inpatients have reduced financial implications. This questionnaire found that the decrease in admissions is marginal and not as significant as anticipated. This could be indicative of an increased hospital due to ADRs, socio-economic circumstances, and ill patients resulting from advanced disease. This also highlights that the decentralization of patients which is anticipated to drastically reduce hospital admissions is not implemented.

Section C of the questionnaire sought to establish aspects relating to the safety and efficacy of the MDR-TB regimens. In terms of the safety of the SBR, the majority of participants (n=12) agreed that there is a strong likelihood of a patient experiencing an ADR. On a severity scale of 1-5 with 1 representing minor and 5 representing severe, 11 participants rated the severity of side effects as 4 which is indicative of severe and life-threatening ADRs. 9 participants rated the severity of the side effects as 3 and 6 participants rated the severity as 5. A grade 3 indicated an ADR of moderate severity and 5 indicates an ADR causing death. The SBR is associated with a better safety profile compared to the kanamycin-based regimen. The SBR is a relatively new regimen therefore there is still a lot that is unknown about the ADR profile (40). The responses from this study indicate that despite the reduced quantity of ADRs, the ADRs are more severe. A study by *Mason et al.* concluded that the use of injectable-free regimens is associated with improved outcomes (40). 23 responses of participants agreed with this conclusion. The reduced number of visits to a hospital or clinic as required by the kanamycin-based regimen was a common response provided. 3 participants found that the simplified regimen enhanced outcomes as it is easier to understand. A minority group (n=5) found that the SBR causes increased loss to follows as patients do not come for appointments and the increased adverse drug reactions encourages defaulting behavior. Converse to literary opinions, the group found that a duration of 9 months impacts negatively on patient outcomes There is data to show that a shorter duration of treatment improves adherence to treatment regimens (40). Contrary to literature that supports the SBR as an effective regimen in reducing loss to follow rates, 7 participants mentioned there was an increase in the number of defaulters associated with the SBR. A minority (n=6) responded that SBR has decreased the number of loss to follow patients and 3 participants found that there was no change in the loss to follow rate. This result however may not be a true reflection of the groups’ opinion as only 47% of participants have answered the question. 13 participants felt that there was an increase in the death rate in patients who used SBR. These deaths may not correlate with the regimen and may be a reflection of late diagnosis, co-morbidity, or poor adherence. The documentation on mortality in the South African setting concludes that the use of bedaquiline in MDR-TB regimens is associated with a decreased mortality compared to regimens such as the kanamycin-based regimen. Initial concerns around the association of bedaquiline with increased mortality were raised from the results of clinical trials which resulted in the WHO exercising caution on the use of bedaquiline (46). Thematic responses to the comparison of the SBR and BPaL regimen highlighted the benefits offered by SBR. The aspects pertaining to ease of administration, reduced loss to follow, and pill burden.

Responses on how beneficial the BPaL regimen will be, found participants at an impasse with both 14 responding that it would be beneficial and 14 responding that it is not the aspect lacking in the TB program. In light of the fact that the participants felt that the SBR played a minor role in loss to follow rates, it can be assumed that those who supported the use of the BPaL did so as the reduced number of drugs coupled with the duration make it a favorable regimen (47). Participants expressed the need for novel regimens to treat MR-TB. High loss to follow rates drive the development of resistance further rendering available medication inadequate. Cross-resistance between bedaquiline and clofazimine further increases the likelihood of XDR-TB. The emergence of resistance to new medication highlights the need for novel regimens such as the BPaL (48). Those participants who did not agree responded that socio-economic factors need to be focused on. The factors included access to health care, education, and understanding of the disease, alcohol, and drug abuse. Two participants mentioned that “drug and alcohol use are detrimental to outcomes of MDR-TB” and “until measures are taken to address the alcohol issues in the country this will always impact compliance”. One participant mentioned that focus should be on TB preventative therapy and TB prevention and education as this would have a greater impact on reducing the incidence of drug-resistant TB.

It was anticipated by 25 participants that the BPaL regimen would be associated with a greater number of severe to life-threatening ADRs. These responses were based on knowledge of two drugs contained in the BPaL regimen.

Participants disagreed that the BPaL regimen will have a greater number of positive outcomes compared to SBR. Thematic analysis of the responses found that more data on safety and efficacy is required, there is an increased number of ADRs are anticipated and the shorter duration may not significantly impact adherence. Two participants mentioned the use of BPaL should be limited as widespread use may promote resistance to novel drugs. The participants who agreed that BPaL is a superior regimen mentioned that the SBR is associated with poor outcomes and also noted that the BPaL regimen may have a lower incidence of ADRs and lower loss to follow.

The sentiment that loss of effective medication occurs through widespread use and abuse of medication is understood by participants. There were 21 responses that BPaL should not be used routinely in MDR-TB. The recommendation is that BPaL should be reserved for cases of XDR-TB or pre-XDR TB.

The policy development framework and guideline development often occur based on expert input and the current evidence. The implementors of the guidelines often have opinions which are not considered. This study provides a voice to those who work directly with patients and provides insight into their experiences on the MDR-TB program. This is especially useful in the guideline development and to establish if their opinions tie in with what literature is reflecting. The lack of reputable literature in South Africa further adds strength to this study and its aims.

### Limitations

The Covid-19 pandemic has had an impact on the study and its outcomes. At the start of the pandemic, there was mobilization of healthcare workers in order to respond to the growing numbers of infected patients with a limited number of human resources. This involved movement of clinical staff specifically nurses and doctors. Priority was given to patients with Covid-19 while all other programs took the proverbial “back seat”. The staff at MDR-TB departments were moved to other departments as admissions for MDR-TB were limited to avoid Covid-19 infections in already compromised patients. At the point of the questionnaire being issued, there were a smaller number of staff than initially anticipated. This limited the sample size and also the generalizability of the results. There were many missing results for open-ended questions as was anticipated. The language barrier may have impacted the interpretation of the questions and therefore impacted the responses. The English language is not the first language for many staff working in the department. The questionnaire was also issued to staff during working hours for completion therefore this required them to participate between work commitments. This could have impacted the uptake and completeness of the questionnaire.

## Conclusion

This study provided an outlook into the perceptions of healthcare workers on MDR-TB program. MDR-TB program is a dynamic one with changing guidelines in response to new information and new drugs. The effective implementation of these guidelines depends on the clinicians who implement these guidelines having a good and thorough understanding of the guideline or updates. The impact of pill burden was highlighted on numerous responses for its impact on loss to follow thereby impacting the outcomes of patients negatively. Factors that were found to impact pill burden were HIV co-infection, socio-economic factors such as lack of finance and support systems. The impact of HIV was re-emphasized as not only a factor driving the TB epidemic but additionally compromising the outcomes of co-infected patients. This study highlighted the influences of socioeconomic factors on the MDR-TB program as this is overlooked in recent years. Much focus has been on regimens and drug development. It is evident from this study that the focus should be on socio-economic factors that negatively impact treatment outcomes and ensuring the availability of safe regimens. More efforts need to be directed at these factors to achieve greater rates of treatment success. In a third-world country, despite having access to effective treatment options the social determinants of health need to be addressed to achieve better control over TB.

Safety was found to be the biggest priority in treating patients. The ADRs also impact outcomes. The most common ADRs are QT prolongation and anemia. This was highlighted as a major ADR associated with the SBR. The safety of MDR-TB regimens was found to most important in designing a regimen. The shorter duration and efficacy were reinforced especially when compared to older regimens. The BPaL regimen despite having the benefits of decreased pill burden and a shorter duration may not be the solution to curb drug-resistant TB in SA. There are concerns over the safety of the regimen particularly the myelosuppression and the possibility of resistance through widespread use.

## Data Availability

All relevant data are within the manuscript and its Supporting Information files.

